# Understanding monogenic Parkinson’s disease at a global scale

**DOI:** 10.1101/2024.03.12.24304154

**Authors:** Johanna Junker, Lara M. Lange, Eva-Juliane Vollstedt, Karisha Roopnarain, Maria Leila M. Doquenia, Azlina Ahmad Annuar, Micol Avenali, Soraya Bardien, Natascha Bahr, Melina Ellis, Caterina Galandra, Thomas Gasser, Peter Heutink, Anastasia Illarionova, Yuliia Kanana, Ignacio J. Keller Sarmiento, Kishore R. Kumar, Shen-Yang Lim, Harutyun Madoev, Ignacio F. Mata, Niccolò E. Mencacci, Mike A. Nalls, Shalini Padmanabhan, Cholpon Shambetova, J Solle, Ai-Huey Tan, Joanne Trinh, Enza Maria Valente, Andrew Singleton, Cornelis Blauwendraat, Katja Lohmann, Zih-Hua Fang, Christine Klein, the Global Parkinson’s Genetics Program (GP2)

**Affiliations:** Institute of Neurogenetics, University of Luebeck, Luebeck, Germany; Department of Neurology, University Clinic Schleswig-Holstein, Luebeck, Germany; Department of Neurology, University of Free State, Bloemfontein, South Africa; Department of Biomedical Science, Faculty of Medicine, University of Malaya, Kuala Lumpur, Malaysia; Department of Brain and Behavioral Sciences, University of Pavia, Pavia, Italy; IRCCS Mondino Foundation, Pavia, Italy; Division of Molecular Biology and Human Genetics, Department of Biomedical Sciences, Faculty of Medicine and Health Sciences, Stellenbosch University, Cape Town, South Africa; South African Medical Research Council, Genomics of Brain Disorders Research Unit, Stellenbosch University, Cape Town, South Africa; Northcott Neuroscience Laboratory, ANZAC Research Institute, Concord, New South Wales, Australia; Faculty of Medicine and Health, University of Sydney, Sydney, NSW, Australia; Department of Molecular Medicine, University of Pavia, Pavia, Italy; Department for Neurodegenerative Diseases, Hertie Institute for Clinical Brain Research, University of Tübingen, Tübingen, Germany; German Center for Neurodegenerative Diseases (DZNE), Tübingen, Germany; Ken and Ruth Davee Department of Neurology and Simpson Querrey Center for Neurogenetics, Northwestern University, Feinberg School of Medicine, Chicago, IL 60611, USA; Translational Neurogenomics, Genomic and Inherited Disease Program, Garvan Institute of Medical Research, Darlinghurst, New South Wales, Australia; Molecular Medicine Laboratory and Neurology Department, Concord Repatriation General Hospital, The University of Sydney, Concord, New South Wales, Australia; Division of Neurology, Department of Medicine, and the Mah Pooi Soo and Tan Chin Nam Centre for Parkinson’s and Related Disorders, Faculty of Medicine, University of Malaya, Kuala Lumpur, Malaysia; Genomic Medicine Institute (GMI), Cleveland Clinic, Cleveland, OH, United States; DataTecnica, Washington DC, USA; Center for Alzheimer’s and Related Dementias (CARD), National Institute on Aging and National Institute of Neurological Disorders and Stroke, National Institutes of Health, Bethesda, MD, USA; Discovery & Translational Research, The Michael J. Fox Foundation for Parkinson’s Research, New York, New York, USA; Department of Clinical Research, Michael J. Fox Foundation for Parkinson’s Research, New York City, NY, USA; Laboratory of Neurogenetics, National Institute on Aging, National Institutes ofHealth, Bethesda, MD, USA

## Abstract

Until recently, about three-quarters of all monogenic Parkinson’s disease (PD) studies were performed in European/White ancestry, thereby severely limiting our insights into genotype-phenotype relationships at global scale. The first systematic approach to embrace monogenic PD worldwide, The Michael J. Fox Foundation Global Monogenic PD (MJFF GMPD) Project, contacted authors of publications reporting individuals carrying pathogenic variants in known PD-causing genes. In contrast, the Global Parkinson’s Genetics Program’s (GP2) Monogenic Network took a different approach by targeting PD centers not yet represented in the medical literature. Here, we describe combining both efforts in a “merger project” resulting in a global monogenic PD cohort with build-up of a sustainable infrastructure to identify the multi-ancestry spectrum of monogenic PD and enable studies of factors modifying penetrance and expression of monogenic PD. This effort demonstrates the value of future research based on team science approaches to generate comprehensive and globally relevant results.

## Introduction

Although monogenic forms of Parkinson’s disease (PD) have been described worldwide, about three-quarters of all PD genetics studies were performed in individuals of European/White ancestry ^1^, formally referred to as Caucasian, thereby severely limiting our current insight into genotype-phenotype relationships at a global, multi-ancestry scale and contributing further to healthcare and research disparities ^2^. The first systematic approach to embrace monogenic PD worldwide, The Michael J. Fox Foundation Global Monogenic PD (MJFF GMPD) project, was built on the MDSGene Database (www.mdsgene.org) that compiles published genotype-phenotype relationships for monogenic PD ^1, 3-5^ and other movement disorders. Corresponding authors of PD-related articles included in MDSGene were contacted and asked to provide additional information on previously reported or newly identified individuals using an electronic case report form ^1, 6^. Data were collected on almost 4,000 individuals from 89 centers in 41 countries, including affected and unaffected carriers of pathogenic variants in genes implicated in monogenic PD ^1^. However, as the focus of this project was to identify established PD research centers with access to and funding for genetic testing, most of the provided information was on individuals of European/White ancestry. The Global Parkinson’s Genetics Program (GP2; https://gp2.org) complements this approach by including all centers with unsolved but suspected monogenic individuals, specifically addressing those that did not have access to genetic testing or that have not participated in PD research before ^7^. The aim is to extend the global network and include centers from formerly underrepresented countries and individuals with diverse ancestry ^8^. The Monogenic Network of GP2 (GP2 MN) focuses on identifying novel monogenic causes of PD but also characterizing genotype-phenotype relations of known monogenic PD at a global scale ^9^. Therefore, the present study builds on the efforts of The MJFF GMPD project, incorporating it into GP2’s MN, and specifically extending the diversity of the current global map of monogenic PD ^8^.

## Methods

Recruitment of collaborating centers for The MJFF GMPD project was based on a systematic online approach to collect individual-level data on individuals with pathogenic variants in known PD genes (*SNCA, LRRK2, VPS35, PRKN, PINK1, PARK7*) and pathogenic or risk variants in *GBA1* ^1^. This included clinical and demographic data to analyze genotype–phenotype relationships ^1^. In contrast, recruitment for the GP2 MN was more inclusive, and initially focused on individuals with an unsolved but suspected monogenic form of PD based on an early age at onset (AAO; ≤50 years of age) and/or a positive family history ^9^. Potential collaborators were additionally identified through personal contacts, participation in PD consortia, and advertising GP2 at congresses. Collaborators from both efforts were divided into three groups based on their involvement in both projects (Group 1), or only in The MJFF GMPD (Group 2) or GP2’s MN (Group 3).

Participants of The MJFF GMPD project received customized emails including relevant information about GP2 and an online survey gauging their interest in participating and their availability of DNA samples. Collaborators interested in joining GP2 were given standardized instructions for the onboarding process to GP2 MN ^9^. Further, all collaborators previously involved in The MJFF GMPD project (Groups 1 and 2) were asked for their permission to merge both efforts and transfer existing data and samples to GP2’s MN. Only centers with GP2 compliance approval until November 2023 are included in this manuscript. Compliance approval is based on an eligible consent form enabling international sample and data sharing and approval by the local Ethics Committee ^9^.

The assignment to underrepresented countries was based on the World Bank’s classification of income status (https://data.worldbank.org/country/XO), classifying countries with low and middle income as underrepresented.

Regardless of available genetic pre-screening, all samples from individuals with eligible clinical data that passed quality control (QC) underwent genotyping with the NeuroBooster Array (NBA), including 1.9 million markers from the Illumina Global Diversity Array and more than 95,000 neurological disease-oriented and population-specific variants, including several hundred known variants in PD-related genes ^10^. Genotyping did not include the analysis of copy number variants (CNV). Detected variants underwent a pathogenicity evaluation based on the criteria of the American College of Medical Genomics (ACMG) ^11^ using Franklin (https://franklin.genoox.com/clinical-db/home), the human genomic variant search engine VarSome ^12^, and MDSGene criteria (https://www.mdsgene.org/methods). Only (likely) pathogenic or known PD risk variants were included. Additionally, variants in *LRRK2* were evaluated based on their impact on the *LRRK2* kinase activity ^13^, while the *GBA1*-PD Browser (https://pdgenetics.shinyapps.io/gba1browser/) ^14^ was used to evaluate identified *GBA1* variants. Efforts to perform short-read whole-genome sequencing including CNV analyses for this cohort are ongoing.

Percentages in the results section are given as valid percentages.

## Data Sharing

GP2 partnered with the online cloud computing platform Accelerating Medicines Partnership - Parkinson’s Disease (AMP PD; https://amp-pd.org) to share data generated by GP2. Anonymized data can be shared upon request and qualified researchers are encouraged to apply for direct access to the data through AMP PD.

## Results

To date, 100 centers from 46 countries (Figure 1) are included in the GP2 MN. All of the 89 centers previously identified through the MJFF GMPD project ^1^ were invited to join the GP2 as part of the ‘merger’, and 38 (42.7 %) of these are now part of the GP2 MN. Reasons for non-participation were diverse, e.g., including no response to the invitation to participate, delay or failure of the onboarding process due to bureaucratic hurdles (e.g., consent forms not meeting the legal requirements of GP2), or inability to provide DNA samples in addition to clinical data.

**Figure 1:**
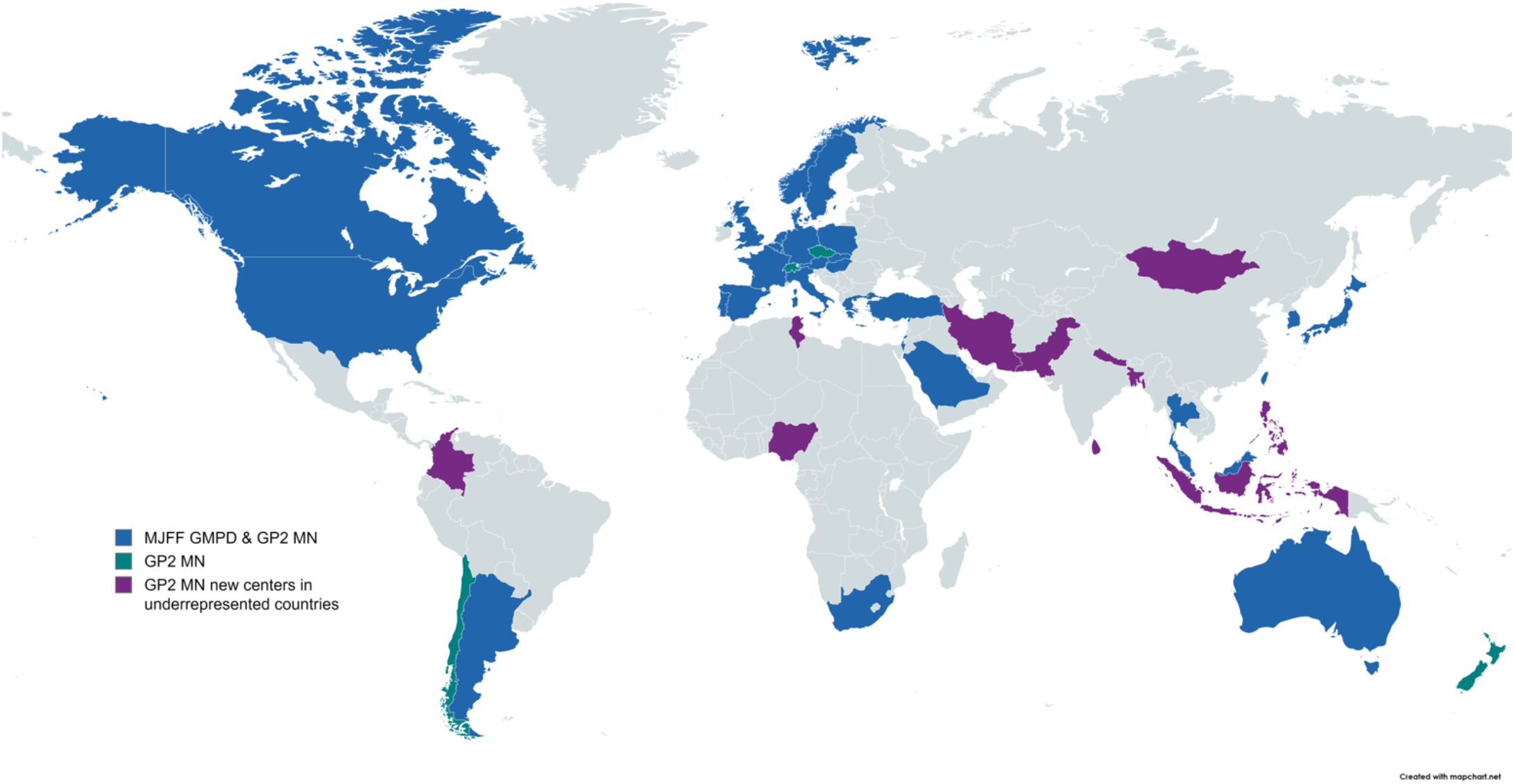
World map of centers participating in the GP2 MN and the MJFF GMPD project. Colored are the countries from which one or more centers participated in the MJFF GMPD project and GP2 MN (blue) or only in GP2 MN (green). Colored in purple are new centers in underrepresented countries that were recruited based on the new approach of GP2 MN. <u>Blue</u>: Argentina, Australia, Austria, Belgium, Canada, Denmark, France, Germany, Greece, Hungary, Israel, Italy, Japan, Luxembourg, Malaysia, Netherlands, Norway, Poland, Portugal, Saudi Arabia, Singapore, Slovakia, South Africa, South Korea, Spain, Sweden, Taiwan, Thailand, Turkey, UK, USA <u>Green</u>: Chile, Czechia, New Zealand, Switzerland <u>Purple</u>: Bangladesh, Colombia, Indonesia, Iran, Mongolia, Nepal, Nigeria, Pakistan, Philippines, Sri Lanka, Tunisia

Nineteen (19%) centers from 15 countries included in the GP2 MN were from underrepresented countries (Figure 1), eleven countries of which were not previously part of The MJFF GMPD project.

Thirty-four centers and the PDGene consortium (https://www.parkinson.org/advancing-research/our-research/pdgeneration) already submitted samples and clinical data to the GP2 MN for further analysis. For 66 centers, the onboarding process is still ongoing. To date, 5,374 DNA or blood samples have been sent to the coordinating GP2 site in Luebeck, Germany, including 468 samples with pending clinical data. The vast majority (n=4,631) of these 4,906 ready-to-analyze samples are from affected individuals with PD/parkinsonism (Table 1), and only a subset (n=275) is from unaffected relatives or unaffected mutation carriers. The median AAO was 49 (Interquartile Range [IQR]: 41-61) years, and 55.7% had an AAO ≤ 50 years (17.5% missing data).

**Table 1:** Clinical Data of the 4,631 individuals with parkinsonism included in the Monogenic Network of the Global Parkinson’s Genetics Program (GP2).

|  | Available data | Missing data<br>n (%) |
| --- | --- | --- |
| **Sex** f = n/m =n | 1899 (42.4%)/ 2582 (57.6%) | 150 (3.2%) |
| **Age** median (IQR) | 66 years (56-75 years) | 295 (6.4%) |
| **AAO** median (IQR) | 49 years (41-61 years) | 809 (17.5%) |
| **Ancestry** | Native Hawaiian/Other Pacific Islander: 1 (<0.1%)<br>American Indian/Alaska Native: 2 (<0.1%)<br>Ashkenazi Jewish: 16 (0.5%)<br>Black/African American: 26 (0.7%)<br>Hispanic/Latino: 70 (2.0%)<br>Arab: 104 (3.0%)<br>White/European: 1471 (41.9%)<br>Asian: 1762 (50.2%)<br>Other: 58 (1.7%) | 1121 (24.2%) |
| **Family history**<br>**of parkinsonism** | Positive: 2103 (59.4%)<br>Negative: 1436 (40.6%) | 1092 (23.6%) |
| **Consanguinity** | Yes: 151 (7%)<br>No: 2017 (93%) | 2463 (53.2%) |

AAO: Age at Onset; IQR: Interquartile range; PD: Parkinson's disease

Genotyping of the GP2 MN cohort with the NBA identified (likely) pathogenic single nucleotide variants (SNVs) in known PD genes (including *GBA1*) in 131 (4.9%) and PD risk variants in *GBA1* and *LRRK2* (*LRRK2* risk variants previously associated with PD in Asian populations) in 272 (10.3%) out of 2,648 individuals tested thus far; the genetic details are listed in Table 2. Heterozygous variants were identified in *GBA1* (n=84), *LRRK2* (n=12), and SNCA (n=14), and one carrier of a pathogenic variant in VPS35 was found. Sixteen carriers of homozygous variants in *PINK1* and 4 *PRKN* homozygous variant carriers were identified but no carriers of biallelic variants in *PARK7* (DJ1). Further, genotyping identified variants of uncertain significance (VUS) in known PD genes in 33 individuals (Table 2).

**Table 2.**
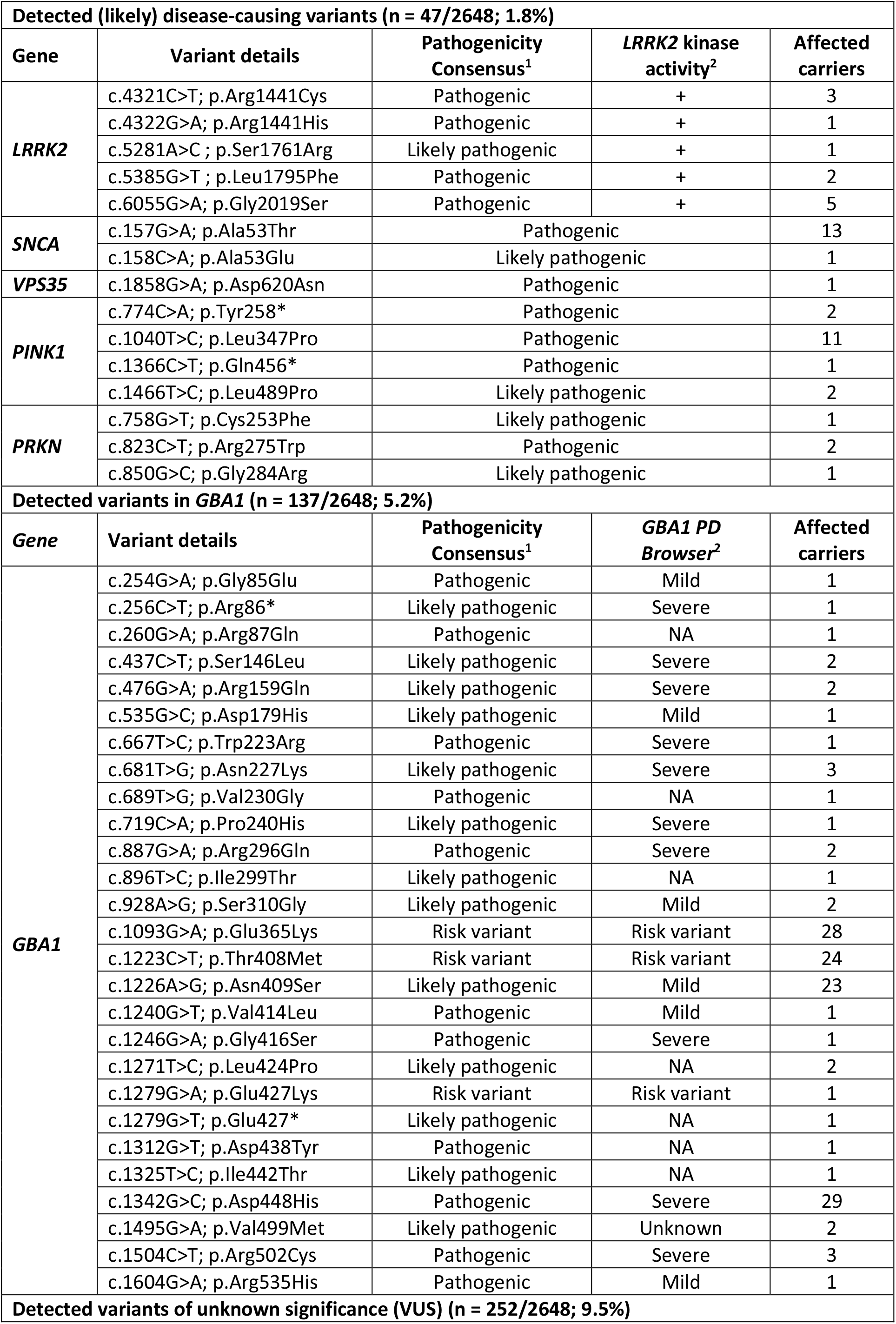
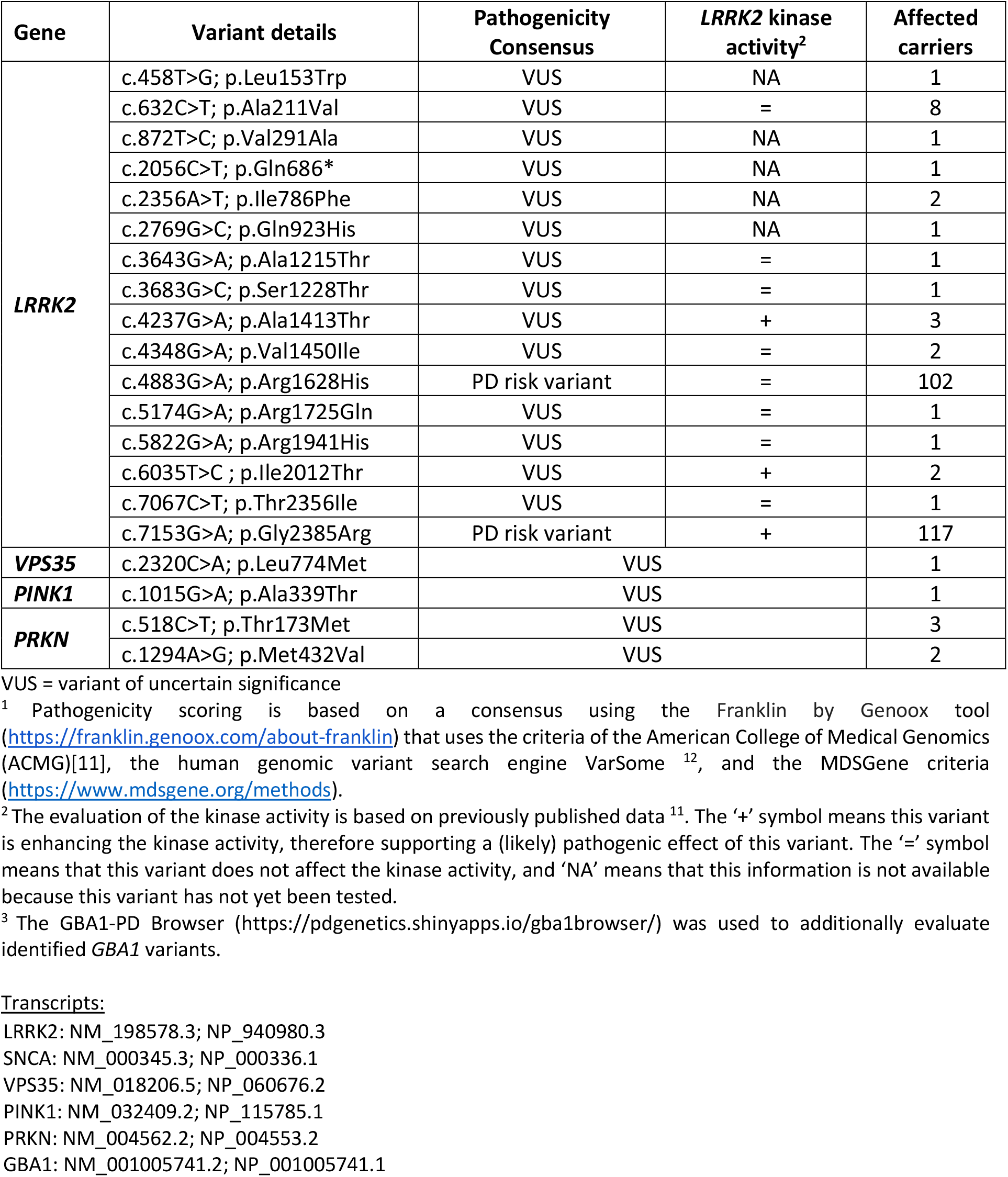
Genetic results from genotyping with the NeuroBooster Array (NBA).

## Discussion

To date, 100 centers from 46 countries are included in the GP2 MN, 38% of which were previously recruited through The MJFF GMPD project ^1^. An important reason for MJFF GMPD centers not to participate in GP MN was that the principal investigator was no longer active, demonstrating the importance of dedicated researchers to drive projects like GP2’s MN. Both projects followed different approaches when recruiting collaborators. The ‘merger’ of both efforts with complementary approaches enables the comprehensive identification of centers working in the PD genetics field, including not only centers identifiable through publications on PD genetics (MJFF GMPD) but also less visible centers (GP2 MN). GP2’s focus on underrepresented regions brought in groups that were not detectable through MJFF GMPD’s publication-based approach, but merging both efforts facilitates the build-up of an inclusive study population with a diverse ancestral background.

Sixty-two centers not previously participating in The MJFF GMPD project were recruited for GP2’s MN, including 16 centers from underrepresented countries. However, the research infrastructure in these countries often posed a challenge to the onboarding process and required intensified efforts and support. Our goal is to overcome these bureaucratic and logistical hurdles to enable global representation of monogenic PD. One way of doing this will be using and supporting established PD consortia in these areas, e.g., LARGE-PD (https://large-pd.org/).

So far, ∼5,400 samples have been included in the GP2 MN and ∼3,900 individuals in The MJFF GMPD project. In The MJFF GMPD project, 3,185 affected and 703 unaffected mutation carriers were included ^1^. Genetic testing of the GP2 MN cohort with the NBA has thus far identified (likely) pathogenic variants in 121 (including *GBA1* variant carriers), PD risk variants in *LRRK2* and *GBA1* in 252, and VUS in 33 out of 2,648 tested individuals. The low frequency of (likely) pathogenic variants compared to previous studies (Westenberger et al., in press) is likely due to the initial selection criteria of GP2’s MN, which prioritized individuals with a negative genetic pre-screening since a major aim of the MN is to discover new PD-related genes ^9^. While GP2’s MN initially focused on unsolved cases, The MJFF GMPD project collected data and samples from individuals with known monogenic causes. Integrating The MJFF GMPD project into GP2 has widened the scope of GP2’s MN. Merging both projects will not only allow creating a sufficient infrastructure to identify the clinical spectrum of monogenic PD across diverse ancestries but also enable investigating factors modifying monogenic PD by identifying a large number of mutation carriers. Together, the combined effort now allows for a more comprehensive, cohesive, and collaborative approach to understanding monogenic PD. Identifying patients with monogenic PD provides the basis for recruiting individuals with genetic PD/parkinsonism for future gene-specific clinical trials which is a dedicated aim of one of the GP2’s special interest groups.

Our efforts of combining two large genetic PD research initiatives underline that the future of research should be based on team science approaches to combine data into even larger, standardized data sets to generate meaningful and globally relevant results ^6^. A collaborative mindset is indispensable to sharing expertise internationally and facilitating new research opportunities which will eventually enable the development of personalized therapies.

## Supporting information

GP2 Banner Author List

## Data Availability

Anonymized data are available upon reasonable request from any qualified investigator to the authors.

## Acknowledgment

Data used in the preparation of this article were obtained from the Global Parkinson’s Genetics Program (GP2). GP2 is funded by the Aligning Science Across Parkinson’s (ASAP) initiative and implemented by The Michael J. Fox Foundation for Parkinson’s Research (https://gp2.org). For a complete list of GP2 members see https://gp2.org.

## Funding Sources

This research was supported in part by the Intramural Research Program of the NIH, National Institute on Aging (NIA), National Institutes of Health, Department of Health and Human Services; project number ZO1 AG000534, as well as the National Institute of Neurological Disorders and Stroke.

GP2 is funded by the Aligning Science Across Parkinson’s (ASAP) initiative and implemented by The Michael J. Fox Foundation for Parkinson’s Research (https://gp2.org).

## Notes

### Competing Interest Statement

The authors have declared no competing interest.

### Author Declarations

There is no overall GP2 Ethics approval. All institutions participating in GP2 have obtained approval from their local Ethics committees which is a prerequisite to be part of the GP2 project. All Ethics approvals and patient information sheets and consent forms were reviewed by GP2's Compliance working group in order to ensure that sample and data sharing is allowed.

### Summary of Updates

Only minor changes were made to the manuscript.

## References

1. Vollstedt EJ, Schaake S, Lohmann K, et al. Embracing Monogenic Parkinson’s Disease: The MJFF Global Genetic PD Cohort. Mov Disord 2023;38(2):286–303.

2. Schiess N, Cataldi R, Okun MS, et al. Six Action Steps to Address Global Disparities in Parkinson Disease: A World Health Organization Priority. JAMA neurology 2022;79(9):929–936.

3. Klein C, Hattori N, Marras C. MDSGene: Closing Data Gaps in Genotype-Phenotype Correlations of Monogenic Parkinson’s Disease. J Parkinsons Dis 2018;8(1):S25–S30.

4. Trinh J, Zeldenrust FMJ, Huang J, et al. Genotype-phenotype relations for the Parkinson’s disease genes SNCA, LRRK2, VPS35: MDSGene systematic review. Mov Disord 2018;33(12):1857–1870.

5. Wittke C, Petkovic S, Dobricic V, et al. Genotype-Phenotype Relations for the Atypical Parkinsonism Genes: MDSGene Systematic Review. Mov Disord 2021;36(7):1499–1510.

6. Vollstedt EJ, Kasten M, Klein C, Group MGGPsDS. Using global team science to identify genetic parkinson’s disease worldwide. Ann Neurol 2019;86(2):153–157.

7. Schumacher-Schuh AF, Bieger A, Okunoye O, et al. Underrepresented Populations in Parkinson’s Genetics Research: Current Landscape and Future Directions. Mov Disord 2022;37(8):1593–1604.

8. Global Parkinson’s Genetics P. GP2: The Global Parkinson’s Genetics Program. Mov Disord 2021;36(4):842–851.

9. Lange LM, Avenali M, Ellis M, et al. Elucidating causative gene variants in hereditary Parkinson’s disease in the Global Parkinson’s Genetics Program (GP2). NPJ Parkinsons Dis 2023;9(1):100.

10. Bandres-Ciga S, Faghri F, Majounie E, et al. NeuroBooster Array: A Genome-Wide Genotyping Platform to Study Neurological Disorders Across Diverse Populations. medRxiv 2023.

11. Richards S, Aziz N, Bale S, et al. Standards and guidelines for the interpretation of sequence variants: a joint consensus recommendation of the American College of Medical Genetics and Genomics and the Association for Molecular Pathology. Genetics in medicine : official journal of the American College of Medical Genetics 2015;17(5):405–424.

12. Kopanos C, Tsiolkas V, Kouris A, et al. VarSome: the human genomic variant search engine. Bioinformatics 2019;35(11):1978–1980.

13. Kalogeropulou AF, Purlyte E, Tonelli F, et al. Impact of 100 LRRK2 variants linked to Parkinson’s disease on kinase activity and microtubule binding. Biochem J 2022;479(17):1759–1783.

14. Parlar SC, Grenn FP, Kim JJ, Baluwendraat C, Gan-Or Z. Classification of GBA1 Variants in Parkinson’s Disease: The GBA1-PD Browser. Mov Disord 2023;38(3):489–495.

